# Changes in diet quality across life transitions from adolescence to early adulthood: a latent growth analysis

**DOI:** 10.1101/2024.02.14.24302819

**Authors:** Yinhua Tao, Melanie Wall, Nicole Larson, Dianne Neumark-Sztainer, Eleanor M Winpenny

## Abstract

**Background:** Adolescence to early adulthood is a period of multiple education-, employment- and family-related life transitions. Changing resources and food environments within the context of these transitions could contribute to significant changes in diet, which persist into later adulthood. This study investigated diet quality trajectories from age 15 to 31 years and changes in diet quality associated with life transitions, by sex.

**Methods:** Data from the Project EAT (Eating and Activity in Teens and Young Adults) study were used to examine diet quality among a longitudinal cohort (n=2,524) across four waves (mean ages of 15, 19, 25 and 31 years). Diet quality was evaluated using the DASH (Dietary Approaches to Stop Hypertension) index. Life transitions were assessed by changes in life circumstances between pairs of waves, including leaving the parental home, leaving full-time education, beginning full-time employment, cohabitating with a partner, and becoming a parent. Average within-person changes in DASH scores were analysed by sex-specific latent growth models, incorporating underlying growth trajectories, five life transitions and baseline socio-demographic characteristics.

**Results:** Both sexes followed a quadratic trajectory of DASH scores, showing decreases in diet quality from waves 1 to 2 followed by increases until wave 4. Compared to females, males had worse diet quality at wave 1, and this sex difference widened at wave 4. Leaving the parental home between waves 1 and 2 was associated with transient decreases in diet quality at wave 2 only for males. For females, cohabitating with a partner and becoming a parent between waves 3 and 4 were respectively related to decreases and increases in diet quality at wave 4. Leaving full-time education and starting full-time employment respectively had long-term negative and positive associations with diet quality for both sexes.

**Conclusions:** Diet quality remained suboptimal throughout adolescence but to some extent improved across early adulthood. A sex-sensitive approach in public health policy is welcome for addressing sex differences in diet quality and dietary changes associated with family-related life transitions. Targeted dietary interventions are beneficial for young people who leave their parental home early or who do not enter into a structured school or workplace environment.

## Introduction

Poor diet quality is a key modifiable risk factor for chronic disease, including diabetes, cardiovascular disease and other non-communicable diseases (NCDs) (Stanaway et al., 2018). A systematic analysis of diet-related health risks in 195 countries found that suboptimal diet, particularly high sodium intake and low fruit and whole grain intakes, contributed to substantial burdens of NCD mortality and morbidity among adults aged 25 years and above (Afshin et al., 2019). However, once individuals enter adult life, dietary patterns are fairly stable and tend to track over time (Craigie et al. 2011; Chong, 2022). The developmental stage of early adulthood may therefore represent a transition from adolescence to adulthood and a window for the development of a high-quality diet to prevent NCD in later life.

Early adulthood (age 16-30 years) is a period of multiple life transitions, which provide the opportunity to disrupt poor eating habits developed during adolescence and to establish healthy dietary patterns persisting into the adulthood (Winpenny et al., 2017; Christoph et al., 2019). Alongside rapid physiological and psychological development (Viner et al., 2015), young people will be exposed to different food-related environments following early adulthood transitions. Specifically, most young people move out of the parental home, which underlies significant changes in the home food environment and allows them more autonomy in food choices (Shepherd and Dennison, 1996; Cruz et al., 2018). They also complete high school education and choose different education/occupational paths, such as continuing further education or starting a first job, leading to changing exposures to institutional food environments. Besides, these life changes are often accompanied with changes in social environment as young people are exposed to a wider social network, develop significant other partner relationships, and may become a parent. A wide range of exposures within these food-related environments are established correlates of quality of diet (Sallis et al., 2015).

There is emerging longitudinal evidence of changes in diet from adolescence to early adulthood. However, these longitudinal studies are not consistent in their approaches to assessing dietary intake or the timing of dietary changes. Studies in the US and Australia showed that diet quality declined or stayed at a low level from age 15 years to the early twenties (Lipsky et al., 2015, 2017; Appannah et al., 2021). In contrast, Winpenny et al. (2018, 2020a) observed a quadratic trajectory in food group consumption for American and Norwegian young people, indicating decreases in diet quality in their early twenties followed by positive changes until the early thirties. Moreover, the timing of early adulthood transitions may be relevant to changes in dietary intake. For example, adolescents who leave full-time education at a younger age (e.g., after high school) are often limited in financial resources and exposed to strong peer networks (Poobalan et al., 2014; Appannah et al., 2020). These factors could lead to significant changes in diet, but it is not clear whether dietary changes associated with life transitions will persist over time. Compared to transient changes in diet following the transitions, persistent dietary changes are more of a concern for public health researchers and practitioners, since tailored interventions are required to prevent unhealthy dietary intake patterns from being carried into adulthood. To address these considerations, longitudinal research needs to take into account the timing of life transitions and differentiate their transient or persistent associations with changes in diet quality, superimposed on underlying dietary trajectories from adolescence to early adulthood.

Another important consideration is sex-related differences in diet and dietary changes across early adulthood. There is evidence of greater excess weight gain among males than females during adolescence (Deforche et al., 2015; Johnson et al., 2015). Less studied is whether males’ excess weight gain is accounted for by their poor diet, and if so, how sex-specific dietary trajectories develop across early adulthood. Given the social roles undertaken, males and females may also change their food consumption in different ways after major transitions in life. A longitudinal study in Norway showed that while young males changed their food intakes with age, females’ intake of food was more responsive to life transitions, particularly leaving the parental home and leaving full-time education (Winpenny et al., 2018).

The current study aims to investigate changes in diet quality and the associations with life transitions from adolescence to early adulthood. To achieve this aim, we analysed longitudinal cohort data from the Project EAT (Eating and Activity in Teens and Young Adults) study to address the following research questions:

1. How does diet quality change from adolescence to early adulthood (age 15 to 31 years)?
2. How are education-, employment- and family-related life transitions associated with changes in diet quality?
3. How do changes in diet quality and associations with life transitions differ by sex?

## Methods

### Study Design and Sample

This study used data from the Project EAT (Eating and Activity in Teens and Young Adults) study, a longitudinal investigation of young people’s eating, activity and weight-related health behaviours (Neumark-Sztainer et al., 2018). The first wave (wave 1) of data was collected in 1998-1999. Participants (mean age 14.9, n= 4746) from 31 public secondary schools in the Minneapolis-St Paul metropolitan area of Minnesota completed the baseline survey, including socio-demographic characteristics, family- and education/employment-related life circumstances, and daily food intake. The follow-up surveys were conducted every five years in 2003-04 (wave 2, mean age = 19.4, n = 2516), 2008-09 (wave 3, mean age = 25.3, n = 2287), and 2015-16 (wave 4, mean age = 31.1, n = 1830). For the current study analysis, longitudinal participants who completed Project EAT surveys and food frequency questionnaires (FFQ) at two or more waves (n = 2524) were included due to the requirement for information on life transitions between pairs of survey waves. Ethical approval for Project EAT was obtained from the University of Minnesota’s Institutional Review Board Human Subjects Committee. Parental consent and written assent from participants were obtained at wave 1. For each follow-up survey wave, participants reviewed a consent form, and completion of the follow-up survey implied written consent.

### Dietary Intake Quality

Dietary intake was measured using two age-appropriate forms of semi-quantitative FFQs. The 1995 version of the Youth and Adolescent Questionnaire was used to assess dietary intake at wave 1 and wave 2, and the 2007 grid form of the Willet Adult FFQ was used at wave 3 and wave 4 (Feskanich et al., 1993; Rockett et al., 1997). The reproducibility, validity, and comparability of the two forms have been described previously (Rimm et al., 1992; Rockett et al., 1995; Larson et al., 2012). A main difference between the two FFQs is that the adolescent form of the FFQ excluded foods that are more commonly consumed during adulthood, and thus included fewer total items than the adult FFQ (127 versus 151 food items). To investigate longitudinal changes in dietary intake, we included food and beverage items included in both adolescent and adult FFQs for the assessment of diet quality.

The DASH (Dietary Approaches to Stop Hypertension) index was used to assess overall diet quality (Fung et al., 2008). This index is based on adherence to a DASH diet, shown to reduce blood pressure and other cardiovascular risk factors in clinical trials (Appel et al., 1997; Siervo et al., 2015). The DASH index recommends a healthy dietary pattern that is rich in the components of fruit, vegetables, whole grains, low-fat dairy, nuts and legumes, and low in the components of red and processed meat, added sugar, and sodium. For each of the components, a score of 0-10 was assigned based on recommended daily servings (Gunther et al. 2009). The DASH index was subsequently calculated by summing the scores of eight components, resulting in a score on a 0-80 scale (0 being the worst and 80 being the best diet quality). Compared to a measure of specific nutrient or food group intake, the DASH index provides a comprehensive assessment of overall dietary intake, taking into account the combination of interrelated nutrients and foods (Miller et al., 2013).

The included food and beverage items for each DASH component and the method for calculation of the DASH index are provided in Table S1. Note that before calculating the DASH index, we excluded the dietary records with implausible energy intakes (< 500 or > 5000 kcal/day; Larson et al., 2012). To assess diet quality independent of reported total dietary intakes, we uniformly adjusted energy intakes to 2000 kcal (8.37 MJ) per day for both sexes using the residual method (Willett, 2012).

### Life Transitions

Five life transitions that are common across early adulthood have been identified using survey measures and previously described in related research (Winpenny et al., 2020a). The transitions included in the analysis are: (1) leaving the parental home, (2) leaving full-time education, (3) beginning full-time employment, (4) cohabitating with a partner, and (5) becoming a parent. These transition variables were assessed by comparing participants’ life circumstances between pairs of waves. The survey questions on life circumstances at each wave were about living arrangements (“During the past year, with whom did you live the majority of the time?”), education status (“Which of the following best describes your student status?”), employment status (“How many hours a week do you work for pay?”), and having children (“How many children do you have?”). For missing data on wave-specific life circumstances, we assumed that life circumstances did not change and thus imputed the data from the previous wave. The transition variables were set as 0 prior to a transition taking place, and 1 following a transition. Considering our interest in life transitions occurring for the first time, we regarded wave-specific life circumstances which returned to the pre-transition state as missing, and thus removed these wave-specific data from the analysis (less than 10% of the participants for each exposure). For example, if a participant left the parental home between waves 1 and 2 and then moved back in together with parents at wave 3, the transition variable of leaving the parental home were set as 0 at wave 1, 1 at wave 2 and as missing at waves 3 and 4.

### Sociodemographic Characteristics

We included baseline socio-demographic characteristics as covariates in our analysis. Note that sex, self-identified as male or female by participants, was not included as a covariate since our analyses were stratified by sex (as elaborated below). The baseline age in years was adjusted for because of the variance in the age of participants when they were recruited at wave 1 (ranging from 11 to 18 years). Race/ethnicity was dichotomised as non-Hispanic white and other ethnicity/race groups. Parental socio-economic status was a five-level composite index, estimated based on parental education levels but also accounting for family eligibility for public assistance, eligibility for free or reduced-cost school meals, and parental employment status (Sherwood et al., 2009). General health status was self-reported by answering the question: “How would you describe your health?”, with four response categories: poor, fair, good, and excellent.

### Statistical analyses

Statistical analyses were conducted in R (version 4.3.1). We compared the socio-demographic characteristics of our study participants and those included in the Project EAT baseline survey, and assessed the occurrence of life transitions and changes in DASH scores across four waves. Independent t-tests were used to examine sex differences in DASH scores and eight DASH components at each wave.

Multiple-group latent growth models were used to estimate the trajectories of DASH scores with age and the associations with life transitions by sex. Before testing sex differences in diet, we constructed the population latent growth model (not differentiated by sex) step by step. First, an unconditional growth model was built to fit the population growth curve of DASH scores, showing that inclusion of a random intercept, random linear slope and random quadratic slope produced the best model fit (Table S2; model 1). Given the recruitment of participants from schools, we tested the school-level variances in DASH scores by calculating intraclass correlation coefficients (ICCs). The results show that ICCs were low (3.76% of total variances) at the school level, suggesting no serious issue from sample clustering by school (Lai and Kwok, 2015), so we did not take account of school-level clustering in our analyses. We next added the baseline covariates as predictors of the three latent parameters.

A final step was the inclusion of life transition variables to test for deviations of the population growth curve after the transitions. We specified two different models to investigate transient and persistent changes in diet quality associated with life transitions. The transient associations were examined by linking each life transition between a pair of waves to the observed DASH score in the following wave (model 2). The persistent associations were analysed by relating each life transition between a pair of waves to an additional latent intercept of DASH scores across the following waves (model 3), interpreted as an overall shift to the underlying growth trajectory (Curran et al., 1998; Winpenny et al., 2020a). To reach a general understanding of the associations between life transitions and diet quality, we additionally fixed the coefficients of each life transition to be the same across four waves in the persistent associations model (model 4). Note that when estimating the associations of the growth parameters with each life transition, we mutually adjusted for the other four life transitions in the models.

After fitting the population growth curve, we tested sex differences in dietary changes in a series of models. This test found that a model where both means and regression parameters varied by sex provided the best fit (Tables S3-4); that is, males and females followed different growth trajectories of diet quality, and the associations of life transitions and covariates with diet quality were also sex-specific. Figure 1 demonstrates the path diagram of sex-specific latent growth models to investigate associations of life transitions with persistent changes in diet quality (model 3). The path diagram of the transient associations model (model 2) is provided in supplementary Figure S1. Throughout the modelling analysis, the maximum likelihood estimator with robust standard errors (MLR) was used to account for the non-normal distribution of DASH scores. Missing data were addressed by using full information maximum likelihood (FIML) estimation. Goodness of fit of the models was assessed based on the comparative fit index (CFI), Tucker–Lewis index (TLI), root mean square error of approximation (RMSEA), standardized root mean square residual (SRMR) and the likelihood-based fit indices.

**Figure 1.**
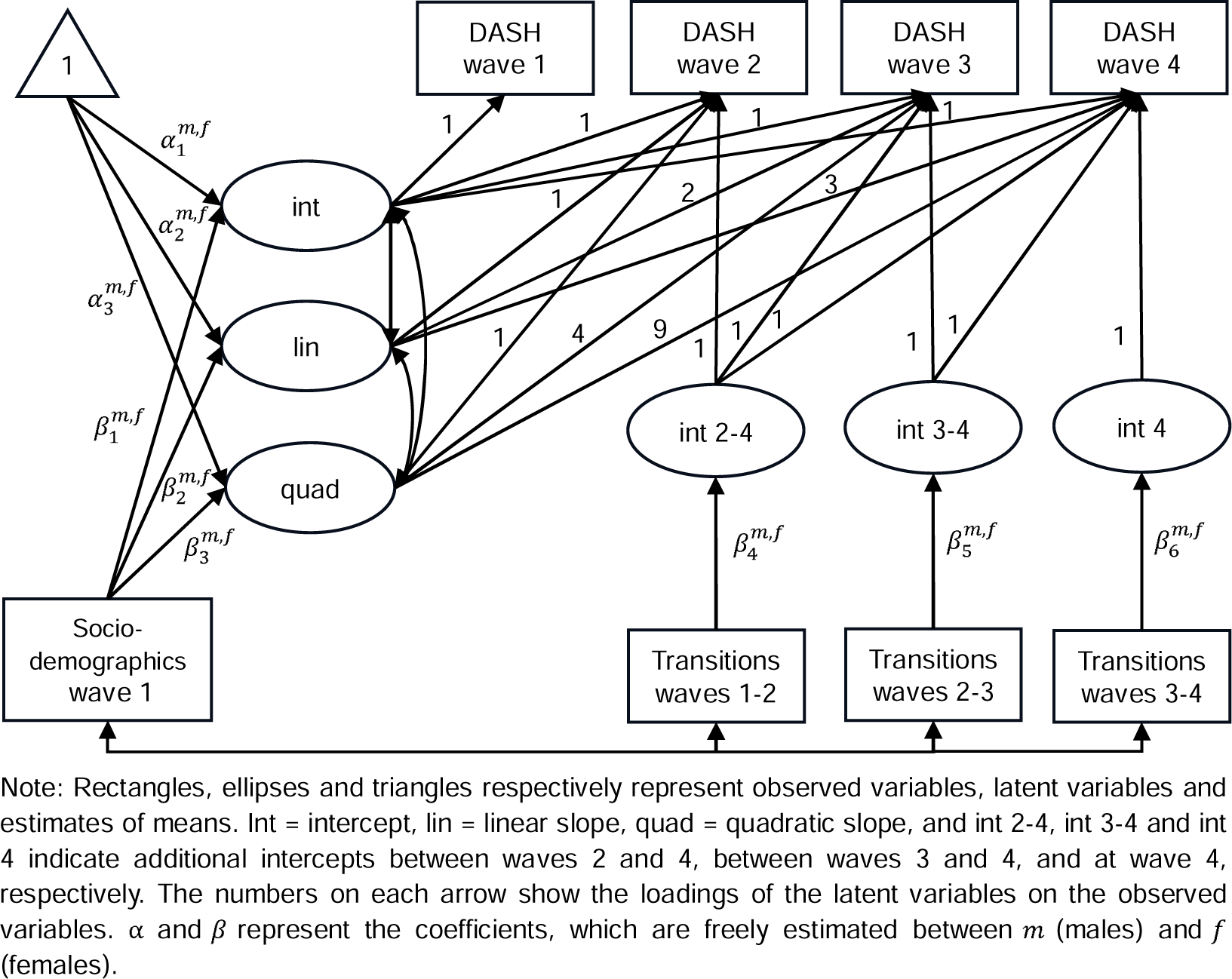
Path diagram of sex-specific latent growth models to investigate associations of life transitions with persistent changes in diet quality after controlling for underlying growth trajectories and baseline socio-demographic characteristics

Figure 2 illustrates the latent growth curves of changes in DASH scores from waves 1 to 4 for males and females, respectively (model 1). The curves for both sexes followed quadratic growth trajectories: predicted DASH scores dropped slightly from waves 1 to 2, and then rose rapidly from waves 2 to 4 (p<0.001 for three latent parameters of males and females). Compared to females, males had lower means for the latent intercept (37.97 versus 40.01) and linear slope (-2.24 versus -0.99). This indicated that males’ diet quality started at a lower level at baseline and had a steeper rate of linear decreases across four waves, resulting in a greater initial decrease in diet quality between waves 1 and 2 and smaller subsequent increases across waves 2 to 4 compared to females. At wave 4, therefore, there was a larger sex difference in predicted DASH scores than those at wave 1.

**Figure 2.**
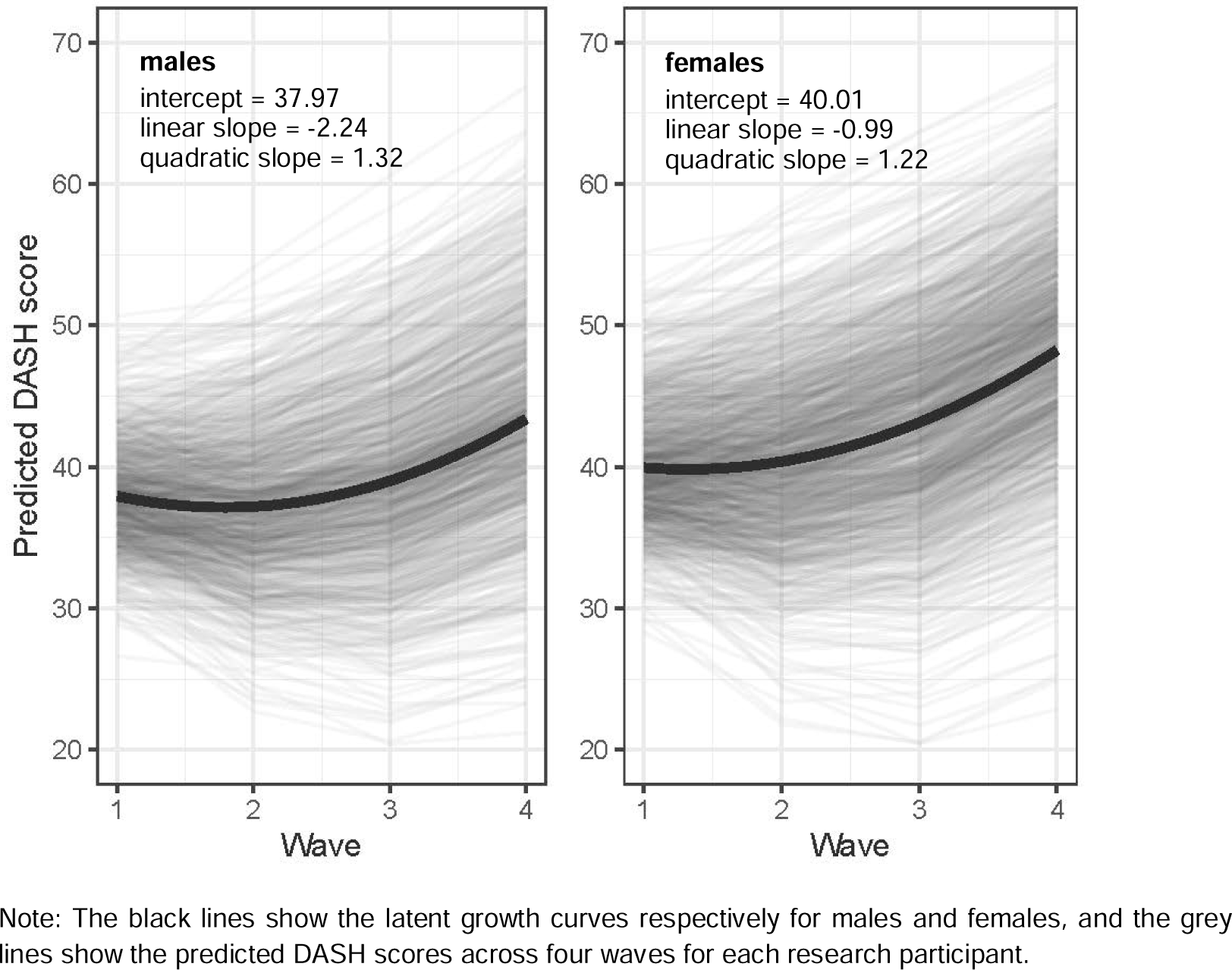
Changes in DASH scores between waves 1 and 4 for males and females, predicted from the unconditional growth model (model 1)

## Results

### Participants’ socio-demographic characteristics

A total of 2542 longitudinal participants who completed FFQs at two or more waves were included in our analyses. At baseline (wave 1), our research participants had a mean age of 14.9 years (standard deviation, SD=1.6), 53.9% self-identified as female, and 74.6% rated their health status as good or excellent. Compared to the original Project EAT participants at baseline, our longitudinal research participants included a higher proportion of non-Hispanic whites (63.6% versus 48.5%) and participants whose parents were of higher socioeconomic status (Table S5).

### Prevalence and timing of life transitions

Across the four survey waves, many research participants experienced each of the five life transitions for the first time between ages 15 and 31 years, ranging from 39.8% of participants becoming a parent to 85.5% leaving full-time education (Table 2). Regarding the timing of each life transition, participants tended to leave their parental home at an earlier age (32.8% of participants between waves 1 and 2 and 30.3% between waves 2 and 3), followed by leaving full-time education (44.3%) and beginning full-time employment (36.8%) occurring most frequently between waves 2 and 3. Cohabitation with a partner, and especially becoming a parent, were more likely to take place in the late twenties between waves 3 and 4. There were few sex differences in the timing and prevalence of life transitions across early adulthood (Table S6). Another notable feature was the parallel changes in life circumstances between pairs of waves. For example, over 60% of participants who left full-time education between waves 1 and 2 (or between waves 2 and 3) also began full-time employment in the same period (Table 2).

**Table 2.**
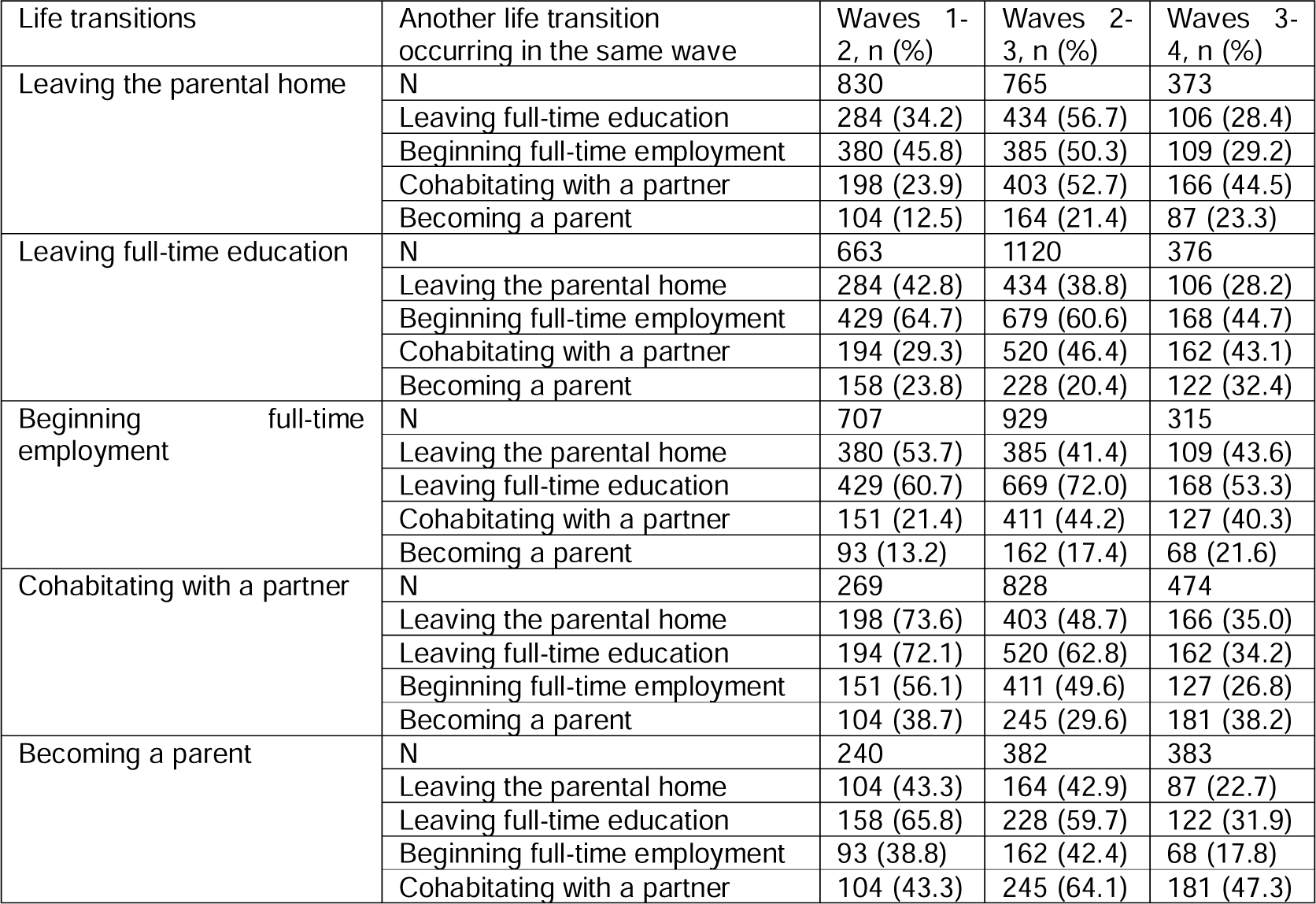
Prevalence and timing of life transitions among research participants (n=2524)

### Changes in diet quality over age for males and females

Compared to participants who identified as female, male participants had significantly worse diet quality in terms of overall DASH scores and most DASH component scores across the four waves (mean overall scores: 36.6-43.5 for males and 39.7-48.3 for females, Table 3). Exceptions were the components of whole grains, low-fat dairy and sodium. While males consumed more daily servings of whole grains and low-fat dairy than females at wave 1, sex differences in the intake of these two healthy foods were negligible in the following three waves, contributing to worse overall diet quality for males in the late period of early adulthood. For the sodium intake, there was little difference by sex across the four waves.

**Table 3.** Descriptive statistics of DASH scores and eight DASH components between waves 1 and 4 for males (n=1163) and females (n=1361)

|  |  | Wave 1 | Wave 2 | Wave 3 | Wave 4 |
| --- | --- | --- | --- | --- | --- |
| DASH scores, mean (SD) | Males | 38.1 (9.4) | 36.6 (9.6) | 40.0 (11.8) | 43.5 (11.8) |
|  | Females | 40.2 (9.3) | 39.7 (9.6) | 44.2 (11.6) | 48.3 (11.0) |
|  | P-value | <0.001 | <0.001 | <0.001 | <0.001 |
| DASH components (servings/day) |  |  |  |  |  |
| Fruit, mean (SD) | Males | 2.2 (2.0) | 1.8 (1.5) | 1.5 (1.3) | 1.6 (1.2) |
|  | Females | 2.8 (2.1) | 2.2 (1.5) | 1.9 (1.6) | 2.1 (1.7) |
|  | P-value | <0.001 | <0.001 | <0.001 | <0.001 |
| Vegetables, mean (SD) | Males | 1.2 (1.1) | 1.2 (1.0) | 1.6 (1.5) | 2.2 (1.8) |
|  | Females | 1.6 (1.2) | 1.7 (1.2) | 2.2 (1.6) | 2.8 (1.9) |
|  | P-value | <0.001 | <0.001 | <0.001 | <0.001 |
| Nuts and legumes, mean (SD) | Males | 0.2 (0.3) | 0.2 (0.4) | 0.3 (0.5) | 0.5 (0.6) |
|  | Females | 0.3 (0.4) | 0.3 (0.4) | 0.4 (0.4) | 0.5 (0.5) |
|  | P-value | 0.008 | 0.009 | 0.01 | 0.22 |
| Whole grains, mean (SD) | Males | 1.0 (1.2) | 1.0 (1.1) | 1.1 (1.3) | 0.9 (0.9) |
|  | Females | 0.8 (0.9) | 1.0 (0.9) | 1.1 (1.0) | 0.9 (0.7) |
|  | P-value | <0.001 | 0.83 | 0.92 | 0.99 |
| Low-fat dairy, mean (SD) | Males | 2.3 (1.8) | 2.1 (1.6) | 1.8 (1.6) | 1.5 (1.3) |
|  | Females | 2.1 (1.6) | 2.0 (1.3) | 1.7 (1.4) | 1.5 (1.2) |
|  | P-value | <0.001 | 0.03 | 0.66 | 0.84 |
| Sodium, mean (SD) | Males | 2.3 (0.4) | 2.3 (0.4) | 2.2 (0.5) | 2.2 (0.5) |
|  | Females | 2.3 (0.4) | 2.4 (0.4) | 2.2 (0.5) | 2.1 (0.5) |
|  | P-value | 0.78 | 0.18 | 0.79 | 0.12 |
| Red and processed meat, mean (SD) | Males | 0.5 (0.4) | 0.5 (0.4) | 0.7 (0.6) | 0.7 (0.5) |
|  | Females | 0.4 (0.3) | 0.4 (0.3) | 0.6 (0.5) | 0.6 (0.5) |
|  | P-value | <0.001 | <0.001 | <0.001 | 0.01 |
| Sugar sweetened beverages, mean (SD) | Males | 1.5 (1.3) | 1.5 (1.3) | 0.9 (1.3) | 0.7 (1.0) |
|  | Females | 1.3 (1.3) | 1.3 (1.2) | 0.7 (1.1) | 0.7 (0.8) |
|  | P-value | 0.01 | <0.001 | <0.001 | <0.001 |

Note: P-values comparing males' and females' diet are calculated based on t values of the independent t-test.

### Growth trajectories of diet quality for males and females

### Associations of life transitions with changes in diet quality for males and females

Tables 4 and 5 respectively show the associations of life transitions with transient (model 2) and persistent (models 3-4) changes in DASH scores for both males and females. Results for the relationship between baseline socio-demographic covariates and diet quality trajectories (for model 4) are provided in Table S7. All associations between a life transition and changes in DASH scores should be interpreted as additional to the underlying growth curve of DASH scores and the influence of other life transitions. Leaving the parental home between waves 1 and 2 was associated with a transient decrease in DASH scores at wave 2 only for males (model 2). However, the negative dietary effect of leaving the parental home between waves 1 and 2 did not persist over time based on its association with the latent intercept of DASH scores across the following three waves (model 3).

**Table 4.** Associations of life transitions with transient changes in DASH scores at the subsequent wave, for males and females.

| | Observed variables of DASH scores at each wave, $\beta$ (95% confidence interval, CI) | | | | | |
| --- | --- | --- | --- | --- | --- | --- |
|  | Model 2 |  |  |  |  |  |
|  | Males |  |  | Females |  |  |
| Life transitions since previous wave | Wave 2 | Wave 3 | Wave 4 | Wave 2 | Wave 3 | Wave 4 |
| Leaving the parental home | -2.32<br>(-3.56, -1.08) | 0.70<br>(-0.93, 2.28) | -0.83<br>(-2.82, 1.16) | -0.62<br>(-1.61, 0.38) | 0.79<br>(-0.36, 1.94) | -1.47<br>(-3.09, 0.15) |
| Leaving full-time education | -2.18<br>(-3.55, -0.82) | -0.38<br>(-1.77, 1.01) | 0.44<br>(-1.61, 2.48) | -1.60<br>(-2.85, -0.35) | -0.78<br>(-1.96, 0.40) | 0.50<br>(-1.09, 2.08) |
| Beginning full-time employment | 0.85<br>(-0.50, 2.20) | 0.93<br>(-0.50, 2.35) | 0.90<br>(-1.42, 3.21) | -0.66<br>(-1.80, 0.49) | 1.86<br>(0.68, 3.04) | 0.85<br>(-0.76, 2.46) |
| Cohabiting with a partner | 0.72<br>(-1.45, 2.89) | 0.68<br>(-0.93, 2.28) | 0.75<br>(-0.95, 2.46) | -0.90<br>(-2.30, 0.51) | -0.12<br>(-1.31, 1.08) | -1.67<br>(-3.15, -0.19) |
| Becoming a parent | -0.01<br>(-2.42, 2.40) | -1.65<br>(-3.83, 0.53) | 0.56<br>(-1.23, 2.34) | 1.10<br>(-0.49, 2.69) | -0.82<br>(2.37, 0.73) | 1.79<br>(0.42, 3.16) |

Note: Models are adjusted for baseline socio-demographic covariates and the underlying growth curve, and are mutually adjusted for other life transitions. Model fit indices for model 1: CFI=0.984, TLI=0.973, RMSEA=0.027, SRMR=0.040.

**Table 5.**
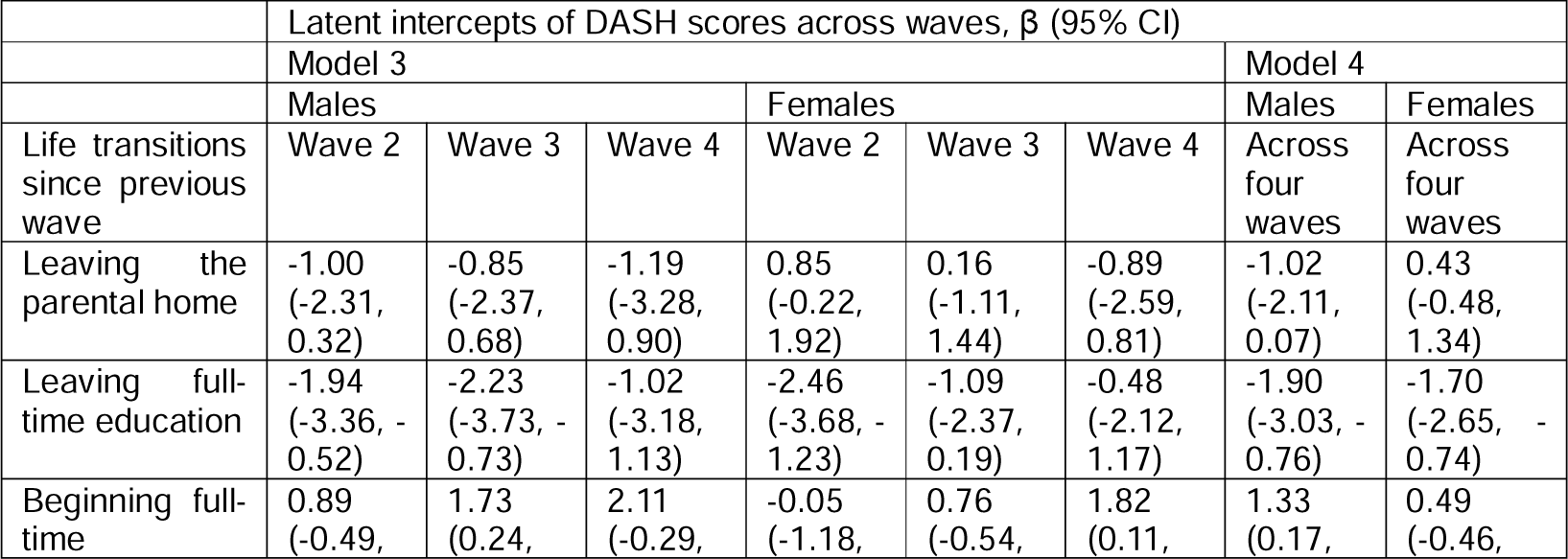
Associations of life transitions with persistent changes in DASH scores across all following waves, for males and females.

Across all waves, leaving full-time education was associated with persistent decreases in DASH scores for both sexes (model 4). When separated by wave, these negative associations were stronger in magnitude when participants left education in earlier waves (models 2 and 3). In contrast, beginning full-time employment showed an overall positive association with long-term diet quality, particularly among males (model 4). In later waves, beginning employment was associated with a larger increase in DASH scores (model 3).

Associations of cohabitation and parenthood with diet quality were observed only among females. Across four waves, females experienced a persistent decrease in DASH scores after cohabitating with a partner (model 4). Between waves 3 and 4, particularly, moving in together with a partner was related to a greater decrease in DASH scores at wave 4 (model 3), thereby reducing the underlying upward growth trajectory of DASH scores in that period. In contrast, after becoming a parent between waves 3 and 4, females increased their DASH scores at wave 4 (model 3).

## Discussion

### Main findings from this study

This longitudinal study enriched the literature on trajectories in dietary intake over the life course by investigating changes in diet quality and associations with life transitions from adolescence to early adulthood. The results show that diet quality slightly decreased from adolescence to the early twenties and then improved until the early thirties for both sexes. Males had worse diet quality scores than females, and this sex difference increased in magnitude across early adulthood. Superimposed on underlying dietary growth trajectories, life transitions in the education/occupational domains were associated with changes in diet quality for both sexes. Leaving early from full-time education (between waves 1 and 2, mean ages 15-19 years) was related to a persistent reduction in diet quality (across waves 2 to 4, mean ages 19-31 years), while starting full-time employment was associated with an improvement in diet quality, with stronger associations observed when this transition occurred in later waves. Sex differences in the associations of diet quality with life transitions were found in the family domain. Leaving the parental home between waves 1 and 2 was associated with a transient decrease in males’ diet quality at wave 2, while females’ diet quality was related to the transitions of cohabitating with a partner (associated with a decrease in diet quality) and becoming a mother (associated with an increase in diet quality), especially when these two transitions took place at a later time of early adulthood (between waves 3 and 4, mean ages 25-31 years).

### Comparison with previous research

Overall diet quality, measured by the DASH index in this study, followed a quadratic growth trajectory from age 15 to 31 years. The observed low diet quality at age 15 (mean DASH score of 39.2) corresponds to the result of a recent systematic review, suggesting that the DASH diet stayed at a suboptimal level during adolescence with the potential to increase the risks of high blood pressure and body mass index gain in the next ten years (Bricarello et al., 2018). Existing longitudinal evidence on changes in overall diet quality from adolescence to early adulthood has shown a stable unhealthy dietary intake, or increasingly worse diet quality until the early twenties (Lipsky et al., 2015, 2017; Smith et al., 2017; Cruz et al., 2018; Appannah et al., 2021). Our analysis extended the evidence by examining longitudinal changes in diet from adolescence to the early thirties, and found a large increase in diet quality (mean DASH scores increasing from 36.6 to 43.5 for males, and from 39.7 to 48.3 for females between waves 2 and 4) after the decrease from adolescence to the early twenties. This finding is consistent with recent studies on the quadratic growth patterns for the intake of specific food groups and nutrients, including fruits, vegetables, saturated fat, sugar-sweetened beverages and fast food, from age 15 to 30 years (Winpenny et al., 2018, 2020a). Another study using the Project EAT data also observed a greater prevalence of participants who met recommendations for daily fruit and vegetable consumption between the mid-twenties and early thirties compared to younger ages (Christoph et al., 2019).

Longitudinal studies on sex differences in dietary trajectories are mixed in their findings for changes in food group consumption from adolescence to early adulthood. The Norwegian Longitudinal Health Behaviour Study reported that the prevalence of daily soft drink consumers was significantly lower among females, although both sexes similarly decreased their intake of fruit and vegetables between ages 14 and 21 years and then increased their intake until the early thirties (Lien et al., 2001; Winpenny et al., 2018). In contrast, the ASH study in the UK showed that despite similar fruit and vegetable intake between both sexes at age 11, females ate more daily servings of fruit and vegetables than males twenty years later (Lake et al., 2009). Some cross-sectional studies suggested that young males had a higher intake of protein, fats and alcohol, while young females consumed fruits, vegetables and wholegrain food, but also sweets and snacks more frequently, making sex differences in overall diet quality less clear (Li et al., 2012; Yahia et al., 2016). To our knowledge, the only evidence of sex-specific growth trajectories of overall diet quality was provided from the Raine Study in Australia (Appannah et al., 2021). Similar to our results, the Raine Study found that males were more likely to follow a trajectory of steady increases in Western diet scores, an indicator of unhealthy dietary patterns, than females from ages 14 to 22 years. Moreover, our results to some extent explained the established evidence of a less healthy diet for males during adulthood (Imamura et al., 2013), which developed from their worse diet quality during adolescence and an increased sex difference across early adulthood.

Few studies have analysed changes in diet quality across life transitions and associated changes in food-related environments, after taking into account long-term dietary growth trajectories. For the transition of leaving the parental home, our results showed that males decreased overall diet quality at wave 2 after moving out of the parental home between waves 1 and 2 (mean ages 15-19 years). This decrease was superimposed on the downward dietary growth trajectory at that time, and thus linked to an additional decline in diet quality. Cross-sectional studies on university freshmen also found that compared to living in the family home, living independently was associated with less healthy eating habits and food intake (e.g., skipping meals more frequently and consuming fewer daily servings of fruit and vegetables; Harker et al., 2010; Pengpid and Peltzer, 2020), especially among male students (Van den Bogerd et al., 2019). The transient decrease in diet quality could result from a sudden change from the home food environment with regular and well-prepared meals by parents. Notably, after specifying the temporal effects of life transitions, our longitudinal analysis indicated that the transient decrease in diet quality after leaving early from parental home did not persist across early adulthood.

Our findings for the transitions of leaving full-time education and beginning full-time employment suggested that spending time in institutional settings, such as schools and workplaces, were beneficial to diet quality across early adulthood. For education-related transitions, there is well-documented evidence of negative changes in weight-related behaviours (e.g., decreases in physical activity and increases in alcohol use) after entering the university, thus contributing to first-semester weight gain (Pullman et al., 2009; Deforche et al., 2015; Hootman et al., 2018). In contrast, little attention has been paid to dietary changes for people who leave education after high school (Poobalan et al., 2014). Our study found that leaving full-time education, especially leaving at an early age, had a long-term association with decreases in diet quality persisting into the early thirties.

For occupational transitions, a recent systematic review showed inconsistent results for changes in food consumption after starting employment (increases in fast food intake but no significant changes in the consumption of fruits, vegetables, confectionery fats and sugar-sweetened beverages; Winpenny et al., 2020b). Our study found that starting a first job showed persistent associations with improvement in diet quality, particularly among males. Possible mechanisms are that workplaces or schools, with a structured day (e.g., days with pre-planned and segmented sections) and a proper place for eating and food purchasing (e.g., dining halls and healthy food stores), help to avoid poor eating habits and improve diet quality (Zosel et al., 2022). Active participation in education and employment activities would also increase people’s income in the long term and thus allow for the purchase of more nutrient-dense foods.

Taking together the results for education- and employment-related life transitions, it is interesting to find that the timing of the transitions mattered for changes in diet quality across early adulthood. For both sexes, leaving early from full-time education was related to a persistent decrease in diet quality. In contrast, the magnitude of the associations between beginning full-time employment and changes in diet quality increased across the waves, such that starting a first job at an older age showed more beneficial dietary changes. As a result, compared to young people who went straight from high-school education into employment, those who continued further education and then entered the job market in the late twenties had better diet quality across early adulthood. This result has important implications for the timing of interventions and the selection of target groups to prevent the establishment of socio-economic inequalities in diet quality during later adulthood (Giskes et al., 2010; Tedstone, 2023).

Family-related transitions of cohabitation and parenthood were associated with changes in diet quality among females, but not among males. Females decreased their diet quality after cohabitating with a partner during the later period of early adulthood. Previous research has found that the associations of cohabitation with diet quality depended on the partner’s own diet (Lake et al. 2009; Berge et al., 2012; Werneck et al., 2020). This conforms to our results considering that males had a worse diet quality than females on average, thereby resulting in a negative impact for cohabitating females and partly cancelling out their underlying increases in diet quality in the late twenties. In addition, there could be a greater social facilitation of eating for females, such as larger serving sizes, less healthy food choices and more sophisticated cooking methods, in cohabitating couples (Burke, 2004; Perry et al., 2016; Smith et al., 2017). For parenthood, previous longitudinal research did not observe significant changes in parents’ intake of selected food groups, nutrients, and overall diet quality (Elstgeest et al., 2012; Laroche et al., 2012; Smith et al., 2017; Corder et al., 2020). After isolating the negative dietary effects of cohabitation from parenthood in the later period of early adulthood, our study found that females experienced an improvement in diet quality after giving birth to a first child in that period. This could result from fewer constraints from financial and time resources at an older age, so that mothers could pay more attention to healthy diet to benefit their children (Burke, 2004).

### Strengths and limitations of this study

This is one of the first longitudinal studies to investigate changes in overall diet quality over a 15-year period from adolescence to early adulthood. Our longitudinal analysis illustrated dietary growth trajectories stratified by sex, and further, examined sex differences in associations of life transitions with transient and persistent changes in diet quality.

This study used a comprehensive measure of diet, the food frequency questionnaire, which allows for assessment of habitual diet, total energy intake and overall diet quality. Project EAT is one of the few studies with longitudinal data on daily intakes of food items and nutrients across early adulthood (Winpenny et al., 2017). However, as with all self-reported measures of diet, these data are subject to participants’ misreporting. In our analyses, we adjusted for total energy intake to estimate diet quality independently from reported energy intake. In addition, any under- or over-reporting of diet quality was unlikely to influence our findings for within-individual dietary changes, unless there were systematic variations in measurement errors with age or across transitions. In this regard, our results for dietary growth trajectories with age could be influenced by secular trends in diet. In the study period of Project EAT; however, other longitudinal studies of different age groups did not find any evidence showing a quadratic growth curve in population dietary patterns (Larson et al., 2016; Christoph et al., 2019).

This study focused on the association of specific life transitions with deviations from sex-specific dietary growth trajectories, after mutually adjusting for other life transitions. This is a much more robust analysis than other longitudinal studies which have examined dietary changes before and after a transition (e.g., Berge et al., 2012; Laroche et al., 2012; Deforche et al., 2015; Smith et al., 2017), without considering other concurrent changes (e.g., another transition in life and changes in diet over age) and the persistence of dietary changes. Our findings therefore provide better causal inference for the effect of a single transition on diet quality in the long term. Even so, our within-individual analysis was unable to investigate how different life transitions interact with each other to (re)shape diet quality over time. For example, the negative dietary effect of leaving the parental home after high school might be modified by following different education- and employment-related life paths, whether they enter into the university education, start their first job, or follow a NEET (not in education, employment, or training) path. A final limitation is that our research participants were mildly overrepresented for non-Hispanic whites and people of high socioeconomic status in the metropolitan area of Minnesota. We welcome future research to validate whether our findings can be generalised to other birth cohorts, populations and regions.

### Conclusions

The age group from adolescence to early adulthood is acknowledged as a difficult group to reach and study (Winpenny et al., 2017). It also represents a critical life stage when a myriad of early adulthood transitions and associated changes in food-related environments take place, which might have a persistent impact on diet quality and diet-related health outcomes in later life. This study found sex differences in dietary growth trajectories in this critical life stage, with males having worse diet quality during late adolescence and smaller increases in diet quality across early adulthood. Moreover, females’ diet quality was more responsive to family-related life transitions, with decreases in diet quality after cohabitating with a partner and increases in diet quality after becoming a parent. For both males and females, spending time in institutional settings, such as schools or workplaces, was associated with better diet quality across early adulthood. Particularly, positive changes in diet quality associated with continued education after high school and beginning employment in a later period of early adulthood could account for well-established socioeconomic inequalities in diet across adulthood. Public health policy and interventions should consider addressing sex differences in dietary changes associated with life transitions in the family domain, and target the population subgroup of early adults who leave the parental home at a young age but do not enter into a structured school or workplace environment.

## Data Availability

The datasets generated and analysed during the current study are not publicly available but are available from the senior author (DN-S) on reasonable request.

## Abbreviations

NCD: Non-communicable disease
EAT: Eating and Activity in Teens and Young Adults
FFQ: Food frequency questionnaires
DASH: Dietary Approaches to Stop Hypertension
ICC: Intraclass correlation coefficient
MLR: Maximum likelihood estimator with robust standard errors
FIML: Full information maximum likelihood
CFI: Comparative fit index
TLI: Tucker–Lewis index
RMSEA: Root mean square error of approximation
SRMR: Standardized root mean square residual
SD: Standard deviation
CI: Confidence interval

## Declarations

### Acknowledgements

The authors would like to thank all participants who contributed data to the Project EAT study, as well as everyone who assisted with the data collection.

### Authors’ contributions

DN-S is principal investigator of the Project EAT study, NL is the Project EAT study project director and managed acquisition of data, YT and EW designed the analyses, YT conducted the analyses and drafted the manuscript, NL, DN-S and EW revised the manuscript, and MW advised on statistical aspects of the study. All authors commented on the analysis plan, contributed to the interpretation of results and critically reviewed the final manuscript. The author(s) read and approved the final manuscript.

### Funding

Data collection for the study was supported by Grant Number R01HL116892 from the National Heart, Lung, and Blood Institute (PI: DN-S). EW and YT were funded by the Medical Research Council MR/T010576/1. The authors’ time to conduct and describe the analysis reported within this manuscript was supported by Grant Number R35HL139853 from the National Heart, Lung, and Blood Institute (PI: DN-S). The content is solely the responsibility of the authors and does not necessarily represent the official views of the National Heart, Lung, and Blood Institute or the National Institutes of Health.

### Availability of data and materials

The datasets generated and/or analysed during the current study are not publicly available but are available from the senior author (DN-S) on reasonable request. All analysis codes are available upon request to the corresponding author (YT).

### Ethics approval and consent to participate

The University of Minnesota’s Institutional Review Board Human Subjects Committee approved all protocols (1207S17861). Parental consent and participant assent obtained at wave 1 and participant consent obtained at each of the subsequent waves.

### Consent for publication

Not required.

### Competing interests

The authors declare that they have no competing interests.

### Rights and permissions

For the purpose of Open Access, the author has applied a Creative Commons Attribution (CC BY) licence to any Author Accepted Manuscript version arising.

## Notes

### Competing Interest Statement

The authors have declared no competing interest.

### Author Declarations

Ethical approval for Project EAT was obtained from the University of Minnesota Institutional Review Board Human Subjects Committee. Parental consent and written assent from participants were obtained at wave 1. For each follow-up survey wave, participants reviewed a consent form, and completion of the follow-up survey implied written consent.

