## Supplementary documents for "Changes in diet quality across life transitions from adolescence to early adulthood: a latent growth analysis"

### Supplementary information

Table S1. Components of the DASH index

| DASH components | Food items | Standard for maximum score of 10 | Standard for minimum score of 0 |
| --- | --- | --- | --- |
| Fruits | Grapes, raisins, bananas, cantaloupe, melons, apples, pears, orange, grapefruit, strawberries, peaches, plums, apricots, orange juice, apple juice or other fruit juice | 0 serving/day | ≥ 4 servings/day |
| Vegetables | Tomatoes, tomato or spaghetti sauce, spinach, greens or kales, peppers, yams or sweet potatoes, zucchini, eggplant, other summer squash, carrots (cooked), carrots (raw), celery, lettuce or iceberg, cabbage or coleslaw, string beans, broccoli, beets, corn, mixed vegetables | 0 serving/day | ≥ 4 servings/day |
| Nuts and legumes | Peanuts, walnuts, other nuts, tofu or other soy protein, beans, lentils or soybeans, peas or lima beans | 0 serving/week | ≥ 4 servings/week |
| Whole grains | Cold breakfast cereal (whole grains), hot breakfast cereal (whole grains), dark bread, other grains like kasha, couscous and bulgur, popcorn | 0 serving/day | ≥ 3 servings/day |
| Low-fat dairy | Skimmed or 1%-2% fat milk, low-fat yogurt, low-fat yogurt (frozen), cottage or ricotta cheese, other low-fat cheese | 0 serving/day | ≥ 2 servings/day |
| Sodium | Sum of sodium content of all foods in FFQs | ≥ 2.57 grams/day | ≤ 2.27 grams/day |
| Red and processed meat | Beef or lamb, pork, hamburger, hotdog, roast beef or ham sandwich, other processed meat sandwich, meatballs or meatloaf, other processed meat | ≥ 4 servings/day | ≤ 2 servings/day |
| Sugar sweetened beverages | Carbonated beverage with sugar, other sugared beverages like sports drink, Punch, sugared ice tea or lemonade | ≥ 1 serving/day | 0 serving/day |

Food intakes are adjusted at 2000 kcal/d. Intakes between maximum and minimum levels are scored proportionally.

Table S2. Fit indices for the intercept, linear and quadratic latent growth models

|  | Intercept | Linear | Quadratic |
| --- | --- | --- | --- |
| Chi-square | 1090.81 | 289.32 | 99.95 |
| Degrees of freedom | 11 | 8 | 4 |
| AIC | 60380.60 | 59535.66 | 59340.85 |
| BIC | 60398.10 | 59570.66 | 59399.19 |
| RMSEA | 0.20 | 0.12 | 0.09 |
| CI for RMSEA | (0.19, 0.21) | (0.11, 0.13) | (0.08, 0.11) |

The model for the cubic setting did not converge due to the four-wave data not allowing for residual errors. AIC: Akaike information criterion, BIC: Bayesian information criterion, RMSEA: Root mean squares error of approximation (RMSEA), and CI: confidence interval.

Table S3. A series of models to test sex differences in the associations of life transitions with transient changes in diet quality

|  | Invariance model | Means model | **Means and regression model** | Means, regression and (co)variances model |
| --- | --- | --- | --- | --- |
| Parameters | 349 | 327 | **300** | 294 |
| -2LL | 1030.34 | 666.79 | **621.73** | 614.47 |
| BIC | 106891 | 106700 | **106867** | 106906 |
| AIC | 105439 | 105119 | **105128** | 105133 |
| $\Delta$ parameters |  | 22 | **27** | 6 |
| $\Delta$ -2LL (p) |  | 363.55 (<0.001) | **45.06 (0.02)** | 7.26 (0.33) |

Invariance model: All parameters are equivalent for both sexes.

Means model: Means of intercepts and slopes differ between both sexes.

Means and regression model: Means of intercepts and slopes, and the regression coefficients of covariates differ between both sexes.

Means, regression and (co)variances model: Means, variances and covariances of intercepts and slopes, as well as the regression coefficients of covariates, differ between both sexes.

Table S4. A series of models to test sex differences in the associations of life transitions with persistent changes in diet quality

|  | Invariance model | Means model | **Means and regression model** | Means, regression and (co)variances model |
| --- | --- | --- | --- | --- |
| Parameters | 346 | 321 | **294** | 288 |
| -2LL | 1019.39 | 658.52 | **610.95** | 603.14 |
| BIC | 106904 | 106739 | **106903** | 106942 |
| AIC | 105434 | 105123 | **105129** | 105134 |
| $\Delta$ parameters |  | 25 | **27** | 6 |
| $\Delta$ -2LL (p) |  | 360.87 (<0.001) | **47.57 (0.01)** | 7.81 (0.28) |

Invariance model: All parameters are equivalent for both sexes.

Means model: Means of intercepts and slopes differ between both sexes.

Means and regression model: Means of intercepts and slopes, and the regression coefficients of covariates differ between both sexes.

Means, regression and (co)variances model: Means, variances and covariances of intercepts and slopes, as well as the regression coefficients of covariates, differ between both sexes.

Tables S3-4 tested sex differences in dietary changes in a series of four models. The first model, the invariance model, assumed all estimated parameters to be invariant between males and females. Estimates from the invariance model were identical to those from the population growth curve model. The second model is the means model, where the means of latent parameters were freely estimated between both sexes. In other words, males and females could follow different growth trajectories of DASH scores with age. The third model, the means and regression model, allowed for sex-specific associations of life transitions and baseline socio-demographics with DASH scores, while the variances and covariances of latent parameters were set free between both sexes in the fourth model, the means, regression and (co)variances model. Results for likelihood-based fit indices of the four models show that the means and regression model provided the best model fit. That is, males and females had different growth trajectories of diet quality, and the associations of life transitions and covariates with dietary growth trajectories were also sex-specific.

DASH

wave 1

DASH

wave 2

DASH

wave 3

DASH

wave 4

Transitions

waves 1-2

Transitions

waves 2-3

Transitions

waves 3-4

Socio-demographics

wave 1

1

1

1

1

3

2

1

1

4

9

1

$$\alpha_{1}^{m, f}$$

$$\alpha_{2}^{m, f}$$

$$\alpha_{3}^{m,f}$$

$$\beta_{1}^{m,f}$$

$$\beta_{2}^{m,f}$$

$$\beta_{3}^{m,f}$$

$$\beta_{4}^{m,f}$$

$$\beta_{5}^{m,f}$$

$$\beta_{6}^{m,f}$$

Note: Rectangles, ellipses and triangles respectively represent observed variables, latent variables and estimates of means. Int = intercept, lin = linear slope, quad = quadratic slope. The numbers on each arrow show the loadings of the latent variables on the observed variables. α and $\beta$ represent the coefficients, which are freely estimated between m (males) and f (females).

Figure S1. Path diagram of sex-specific latent growth models to investigate associations of life transitions with transient changes in diet quality after controlling for underlying growth trajectories and baseline socio-demographic characteristics

Table S5. Baseline socio-demographic characteristics of the participants included in this study and the original Project EAT participants at wave 1

| Socio-demographic characteristics | Participants included in this study, n=2524 | Original Project EAT participants at wave 1, n=4746 |
| --- | --- | --- |
| Age at wave 1, mean (SD) | 14.9 (1.6) | 14.9 (1.7) |
| Sex: female-identified, % (n) | 53.9 (1361) | 49.8 (2357) |
| Race/ethnicity: non-Hispanic white, % (n) | 63.6 (1590) | 48.5 (2264) |
| Parental socioeconomic status, % (n) |  |  |
| 1 (the lowest) | 12.5 (310) | 17.4 (793) |
| 2 | 16.4 (406) | 18.8 (857) |
| 3 | 25.5 (633) | 26.6 (1209) |
| 4 | 28.4 (705) | 23.4 (1065) |
| 5 (the highest) | 17.2 (425) | 13.8 (626) |
| Health status: good or excellent, % (n) | 74.6 (1793) | 73.6 (3195) |

Table S6. Prevalence and timing of life transitions for males (n=1163) and females (n=1361)

|  |  | Males, n (%) | | | Females, n (%) | | |
| --- | --- | --- | --- | --- | --- | --- | --- |
| Life transition | Another life transition  occurring in the same wave | Waves 1-2 | Waves 2-3 | Waves 3-4 | Waves 1-2 | Waves 2-3 | Waves 3-4 |
| Leaving the parental home | n | 353 | 354 | 160 | 477 | 411 | 213 |
|  | Leaving full-time education | 115 (32.6) | 192 (54.2) | 39 (24.4) | 169 (35.4) | 242 (58.9) | 67 (31.5) |
|  | Beginning full-time employment | 178 (50.4) | 181 (51.1) | 51 (31.9) | 202 (42.3) | 204 (49.6) | 58 (27.2) |
|  | Cohabitating with a partner | 65 (18.4) | 169 (47.7) | 77 (48.1) | 133 (27.9) | 234 (56.9) | 89 (41.8) |
|  | Becoming a parent | 28 (7.9) | 69 (19.5) | 33 (20.6) | 76 (15.9) | 95 (23.1) | 54 (25.4) |
| Leaving full-time education | n | 312 | 513 | 156 | 351 | 607 | 220 |
|  | Leaving the parental home | 115 (36.9) | 192 (37.4) | 39 (25.0) | 169 (48.1) | 242 (39.9) | 67 (30.5) |
|  | Beginning full-time employment | 213 (68.3) | 309 (60.2) | 78 (50.0) | 216 (61.5) | 370 (61.0) | 90 (40.9) |
|  | Cohabitating with a partner | 66 (21.2) | 209 (40.7) | 70 (44.9) | 128 (36.5) | 311 (51.2) | 92 (41.8) |
|  | Becoming a parent | 49 (15.7) | 78 (15.2) | 48 (30.8) | 109 (31.1) | 150 (24.7) | 74 (33.6) |
| Beginning full-time employment | n | 345 | 414 | 147 | 362 | 515 | 168 |
|  | Leaving the parental home | 178 (51.6) | 181 (43.7) | 51 (34.7) | 202 (55.8) | 204 (39.6) | 58 (34.5) |
|  | Leaving full-time education | 213 (61.7) | 309 (74.6) | 78 (53.1) | 216 (59.7) | 370 (71.8) | 90 (53.6) |
|  | Cohabitating with a partner | 61 (17.7) | 172 (41.5) | 67 (45.6) | 90 (24.9) | 239 (46.4) | 60 (35.7) |
|  | Becoming a parent | 38 (11.0) | 69 (16.7) | 33 (22.4) | 55 (15.2) | 93 (18.1) | 35 (20.8) |
| Cohabitating with a partner | n | 87 | 332 | 228 | 182 | 496 | 246 |
|  | Leaving the parental home | 65 (74.7) | 169 (50.9) | 77 (33.8) | 133 (73.1) | 234 (47.2) | 89 (36.2) |
|  | Leaving full-time education | 66 (75.9) | 209 (63.0) | 70 (30.7) | 128 (49.5) | 311 (48.2) | 92 (24.4) |
|  | Beginning full-time employment | 61 (70.1) | 172 (51.8) | 67 (29.4) | 90 (49.5) | 239 (48.2) | 60 (24.4) |
|  | Becoming a parent | 32 (36.8) | 101 (30.4) | 88 (38.6) | 72 (39.6) | 144 (29.0) | 93 (37.8) |
| Becoming a parent | n | 72 | 148 | 209 | 168 | 234 | 275 |
|  | Leaving the parental home | 28 (38.9) | 69 (46.6) | 33 (15.8) | 76 (45.2) | 95 (40.6) | 54 (19.6) |
|  | Leaving full-time education | 49 (68.1) | 78 (52.7) | 48 (23.0) | 109 (64.9) | 150 (64.1) | 74 (26.9) |
|  | Beginning full-time employment | 38 (52.8) | 69 (46.6) | 33 (15.8) | 55 (32.7) | 93 (39.7) | 35 (12.7) |
|  | Cohabitating with a partner | 32 (44.4) | 101 (68.2) | 88 (42.1) | 72 (42.9) | 144 (61.5) | 93 (33.8) |

Table S7. Associations of baseline socio-demographic covariates with latent growth parameters in the persistent associations model (model 4)

|  | Latent growth parameters of DASH scores, β (95% CI) | | | | | |
| --- | --- | --- | --- | --- | --- | --- |
|  | Males | | | Females | | |
|  | Intercept | Linear slope | Quadratic slope | Intercept | Linear slope | Quadratic slope |
| Age at wave 1 | **-0.51 (-0.85, -0.17)** | 0.24 (-0.38, 0.86) | 0.01 (-0.19, 0.21) | -0.03 (0.34, 0.29) | 0.50 (-0.04, 1.04) | **-0.18 (-0.35, -0.01)** |
| Race/ethnicity: non-white  (reference: white) | **2.03 (0.81, 3.26)** | **-2.23 (-4.28, -0.17)** | 0.19 (-0.52, 0.90) | **1.79 (0.68, 2.90)** | -1.35 (-3.00, 0.30) | -0.02 (-0.55, 0.53) |
| Parental socio-economic status | **1.19 (0.73, 1.66)** | -0.01 (-0.76, 0.73) | 0.10 (-0.15, 0.34) | **1.10 (0.69, 1.51)** | 0.13 (-0.49, 0.75) | -0.06 (-0.27, 0.15) |
| General health status | 0.67 (-0.13, 1.47) | -0.38 (-1.57, 0.80) | 0.08 (-0.32, 0.48) | **2.09 (1.34, 2.84)** | -0.10 (-1.25, 1.05) | 0.01 (-0.36, 0.38) |
